# The impact of integrating oral health education in cardiac rehabilitation: A randomised controlled trial

**DOI:** 10.1101/2025.11.03.25339448

**Authors:** Lauren. A. Church, Shalinie King, Simone Marschner, Robert Zecchin, Nikki Barrett, Tanya Kumar, Axel Spahr, Clara. K. Chow

**Author notes:** Corresponding author: Address: Sydney Dental School, Block K, Level 6, 176 Hawkesbury Road, Westmead NSW Australia 2145 (LC).

## Abstract

**Background:** Despite the growing body of evidence supporting an association between oral disease and cardiovascular disease (CVD), oral health as a modifiable risk factor for cardiovascular health is rarely addressed.

**Objective:** To determine whether a novel oral health education program improves oral health related behaviours and knowledge of individuals attending cardiac rehabilitation.

**Methods:** This randomised clinical trial (1:1:1) in Australian public hospitals compared three interventions: Group A) individualised oral hygiene instruction (OHI) combined with a digital oral health education (DOHE) package, Group B) DOHE alone, or Group C) usual care: no oral health education. All participants received OHI+DOHE after final follow-up. The primary outcome was improvement in oral hygiene, assessed by approximal plaque index (API) at 6 weeks. Secondary outcomes included API reductions at 12 weeks, as well as self-reported changes in oral hygiene behaviours, knowledge, confidence, status, and motivation.

**Results:** A total of 158 participants were randomised (mean age 62 ± 11 years; 82% males). At six weeks, 77.1% (27/35) of participants receiving OHI+DOHE showed a reduction in API, compared to 26.8% (11/51) receiving usual care (odds ratio [OR] 9.16, 95% CI: 3.34-27.75). Participants receiving DOHE alone showed similar reductions.

**Conclusion:** Oral health education delivered during cardiac rehabilitation using digital media alone, or in combination with clinician delivered face-to-face messaging improves oral hygiene, behaviours and knowledge among patients with CVD and can be integrated into cardiology care.

**Trial registration:** anzctr.org.au ACTRN12623000449639p

## Introduction

The impact of modifiable risk factors such as diet, exercise, alcohol intake, and lifestyle habits on cardiovascular disease (CVD) are well established.^1,2^ However, despite the growing body of evidence linking oral diseases such as periodontitis and tooth loss to the increased risk of CVD,^3–5^ oral health as a modifiable risk factor for CVD is not a routine component of cardiovascular health care.^1,2^

Periodontitis is a multifactorial inflammatory condition^6^ initiated by inadequate oral hygiene, leading to the accumulation of oral biofilm on oral tissues.^7^ The immune response to this biofilm triggers vasodilation of gingival tissues,^6^ facilitating the translocation of bacteria and their products via the bloodstream. Continued absence of oral hygiene increases the local inflammatory response and results in the release of circulating inflammatory markers, including high-sensitive C-reactive protein and interlukin-6.^8^ These markers are strongly associated with an increased risk of CVD, primarily atherosclerosis.^9^ Untreated, periodontitis can lead to tooth loss which therefore is a reflection of an individual’s burden of oral disease.^10^

Good oral hygiene practices including toothbrushing twice daily and cleaning between teeth once daily can prevent bacteraemia and systemic inflammation initiated by the accumulation of oral bio film.^11^ Oral health education integrated into CVD management could be an approach to promote oral hygiene and reduce the inflammatory burden.^12^ Barriers such as cost and long waiting times for public dental services limit access to oral health care, with nearly half of all Australians not visiting an oral health practitioner in 2021 and 2022.^13^ As a result, many individuals do not receive oral health education that could reduce the burden of oral disease. Patients in cardiovascular rehabilitation centres, who are at high risk for future cardiovascular events, may have the motivation to focus on their health following a recent cardiovascular event.^14^ This study aimed to evaluate the efficacy and feasibility of both traditional and digital oral health education in improving oral hygiene as measured by the approximal plaque index (API); and self-reported oral health related behaviours, knowledge, confidence in self-care, oral health status and motivation among individuals attending cardiac rehabilitation services in Australia.

## Methods

The trial was approved by the Western Sydney Local Health District Human Research Ethics Committee: 2023/ETH00516 and registered with Australian New Zealand Clinical Trials Registry: ACTRN12623000449639p. A validated questionnaire tool^15^ adapted for this study was used to collect study data. The trial protocol has been published elsewhere.^16^ This report follows the Consolidated Standards of Reporting Trials (CONSORT) reporting guideline for randomised clinical trials (Supplementary File 1).

### Study design

A randomised, dual centre, parallel design, three arm, single blind, clinical trial of patients attending Cardiac Education and Assessment Programs (CEAP) in Western Sydney Hospitals.

### Population

Eligible patients were aged 18 years or over, with ≥ 1 tooth, approximal plaque index (API) score of ≥60% reflecting poor oral hygiene and attending CEAP due to CVD. Patients were recruited from Westmead Hospital and Blacktown Mt Druitt Hospital, located in Western Sydney; serving diverse populations (46.8% born overseas).^17^ Participants were excluded if they had heart failure, or had/were waiting for a heart transplant; were unable to read or understand English to provide written informed consent, had visited an oral health practitioner for treatment of periodontal disease or received a professional dental clean within the last 6 months.

### Recruitment and consent

One clinical examiner (a trained and calibrated oral health therapist) screened all patient files and approached eligible participants in-person prior to, or after their CEAP appointment. Written consent was obtained via eConsent forms within REDCap^18,19^ on institutional iPads.

### Randomisation and Masking

Research assistants, who were final year oral health or dentistry students, independently randomised participants using the randomisation allocation table within REDCap. Randomisation was 1:1:1 ratio into one of 3 arms of the study: Group A) individualised oral hygiene instruction (OHI) combined with a digital oral health education (DOHE) package, Group B) DOHE alone, or Group C) usual care/control. The clinical examiner left before randomisation, and the proceeding intervention were completed to ensure blinding to the treatment allocation. Participants were not able to be blinded due to the nature of the interventions.

## Interventions

In brief, the same research assistants provided individualised OHI with face-to-face instructions, following a pre-determined standard protocol^16^ at baseline, 6- and 12- week follow-up, and if not possible to attend, via telephone. Demonstrations of proper brushing and interdental cleaning were reinforced on dental models.

Videos included in the DOHE package were chosen based on feedback from a non-dental panel consisting of seven clinicians and non-clinicians. Videos were assessed on specified criteria via anonymous REDCap surveys^18,19^ (Supplementary File 2). Six videos with the highest votes were selected covering a range of topics including the link between oral health and CVD, as well as oral health behaviours. The duration of each video ranged from 1- to 2.19 minutes in length (see Table S1 Supplementary File 3 for video content and timing). At the end of the study, all participants had access to all components of the interventions.

### Trial Procedures

Participants underwent assessments at baseline, 6-weeks and 12-weeks (±2-weeks). Demographic information, medical history, and anthropic measurements were collected at baseline using participant medical files and self-reported data.

Clinical assessments, along with questionnaire data on oral hygiene behaviours, confidence, oral health status, motivation, and knowledge were collected at each time point (Supplementary File 3), before participants received their intervention. Participants unable to attend their follow-up clinical assessments were contacted by phone to complete questionnaires, received OHI and access the DOHE package which was sent via a link.

### Trial outcomes

The primary outcome was whether or not there was a reduction in API (a dichotomous measure of oral biofilm and surrogate measure of oral hygiene) between baseline and 6-weeks. An API reduction was determined as any reduction at follow-up compared to baseline. The same clinical examiner performed all clinical measurements including API, sulcus bleeding index (SBI); a dichotomous measure of bleeding and surrogate measure of gingival inflammation, at each visit, as well as a periodontal screening and recording (PSR), a measure of periodontal treatment needs at baseline (Tables S2 & S3 Supplementary File 3).

Secondary outcomes included self-reported changes in oral health behaviours, knowledge, confidence, status, and motivation. Knowledge questions consisted of 12-items categorised into three categories: impact of oral health on heart health, heart medication implications, and oral disease knowledge. Percentages were calculated to show how many in each study arm answered every question correctly within each category. Questionnaires are in Supplementary File 4.

Participants were considered as changing their behaviour if they answered ‘yes’ to making a change at follow-up. Participants were determined to be confident in oral self-care by answering strongly agree or agree to three oral self-care questions: knowing how to look after their teeth, when they had a dental problem, and when to seek dental treatment. They had good oral health status if rated it as either good, very good or excellent. They were deemed regular attenders if they reported 3 or 6 monthly visits at baseline. They were motivated to visit an oral health practitioner if they answered ‘yes-I have made an appointment’ or ‘yes – I have not yet made an appointment’ at follow-up. Participants provided feedback on each video’s understanding, likeability, and educational value using an online survey where each question was scored on a 5-point Likert scale.

### Data analysis

A pre-specified statistical analysis plan was used according to the intention-to-treat principle. The primary outcome of API reduction or not was assessed using logistic regression to calculate the odds ratio (OR) and their 95% confidence intervals (CIs) adjusting for age (≤64 and ≥65 years) and site (Westmead and Blacktown Mt Druitt). Secondary outcomes were assessed using logistic (categorical variables) or linear regression (continuous variables). The sample size of 165 (55 participants per arm) was calculated to have 80% power (2-tailed, type 1 error 5%) to detect a difference in API reduction between OHI+DOHE and usual care, allowing for a 10% attrition rate. To address missing data, two sensitivity analyses were conducted using the worst-case scenario method and the multiple imputation by chained equations (MICE) method.^20^ The significance level for treatment comparisons for all outcomes was 0.05. All analyses was performed using R version 4.4.2 (R Core Team 2024).^21^

### Results Characteristics

A total of 158 participants were recruited from Westmead (56%) and Blacktown Mt Druitt (44%) CEAP between August 22^nd^, 2023, and 29^th^ August 2024. The recruitment target was not met due to insufficient research assistant coverage. At 6-weeks, a total of 117 participants returned for the in-person clinical exam, and an additional 15 completed the questionnaire only. At 12-weeks, 55 returned for the clinical exam, with an additional 17 completing the questionnaire (Figure 1).

**Figure 1.**
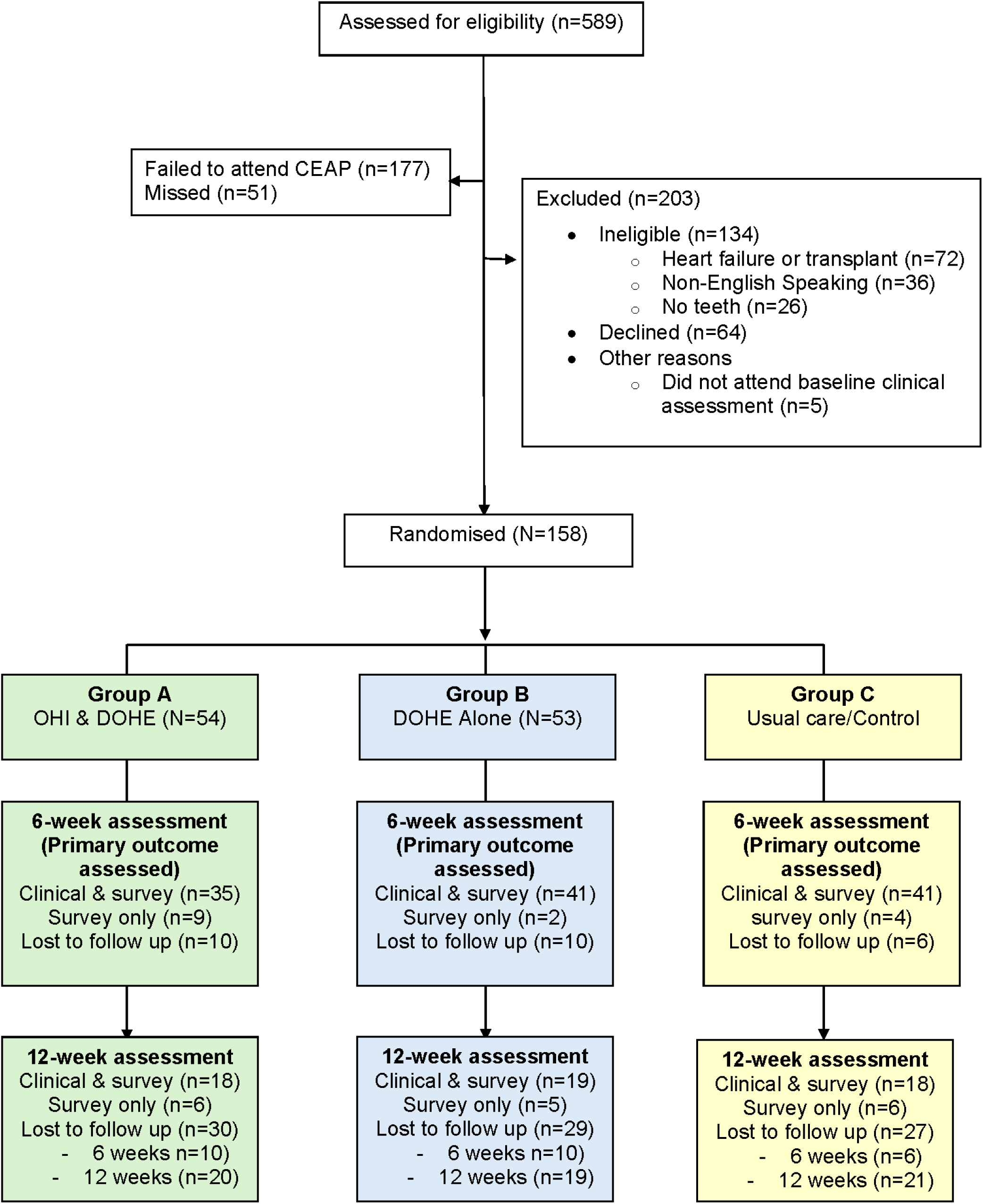
CONSORT diagram.

The mean age of participants was 62±11 years, with 82% male and 40% of participants identifying as Asian (South, East, South-East). All participants had CVD, with 92% having an atherosclerotic aetiology. One or more comorbidities were reported by 71%, with hypertension the most prevalent (Table 1). Just under six percent were current smokers and 8.9% were daily drinkers. Only 5.1% of participants received oral health advice after their cardiac diagnosis.

**Table 1.**
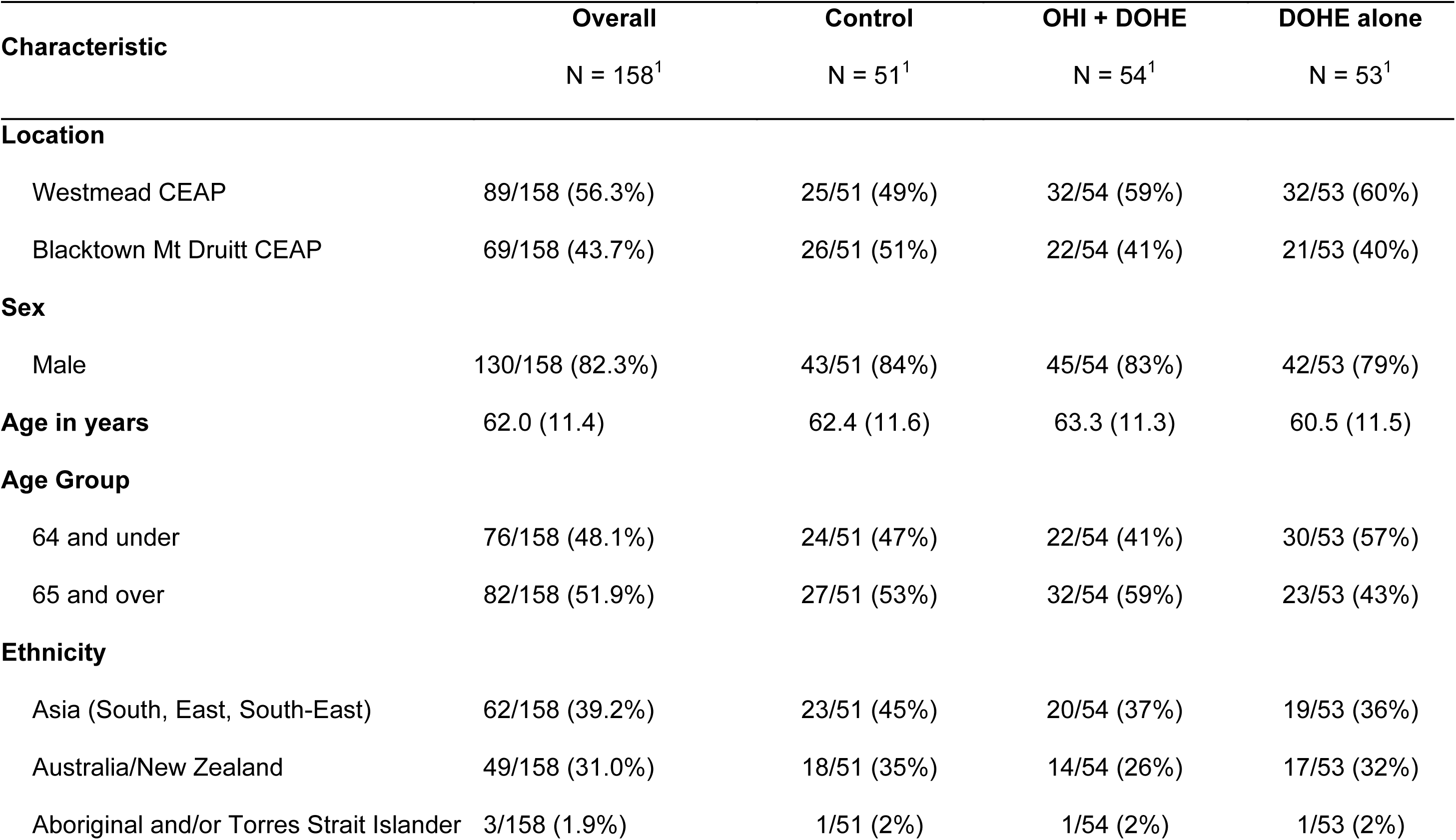
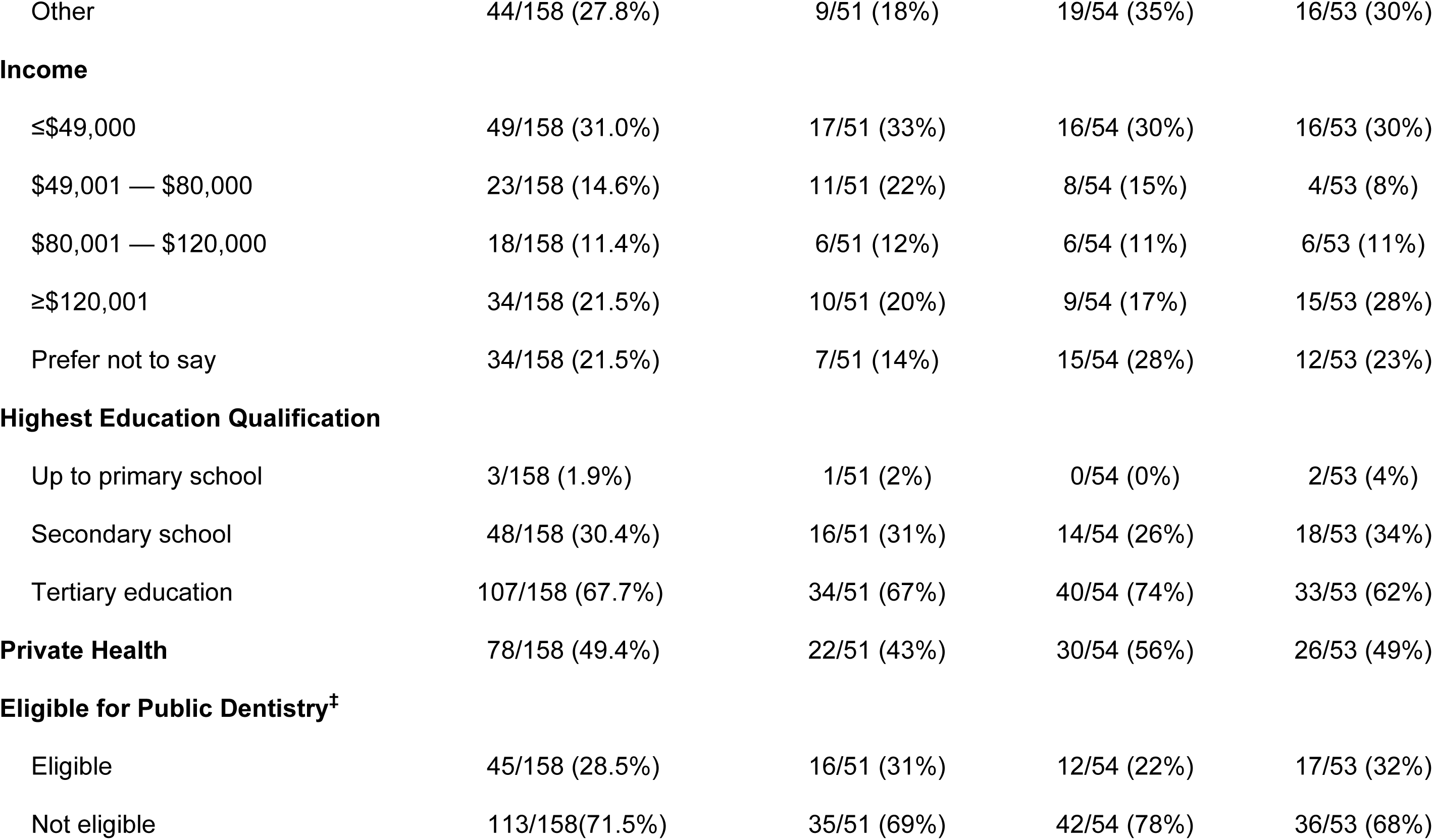
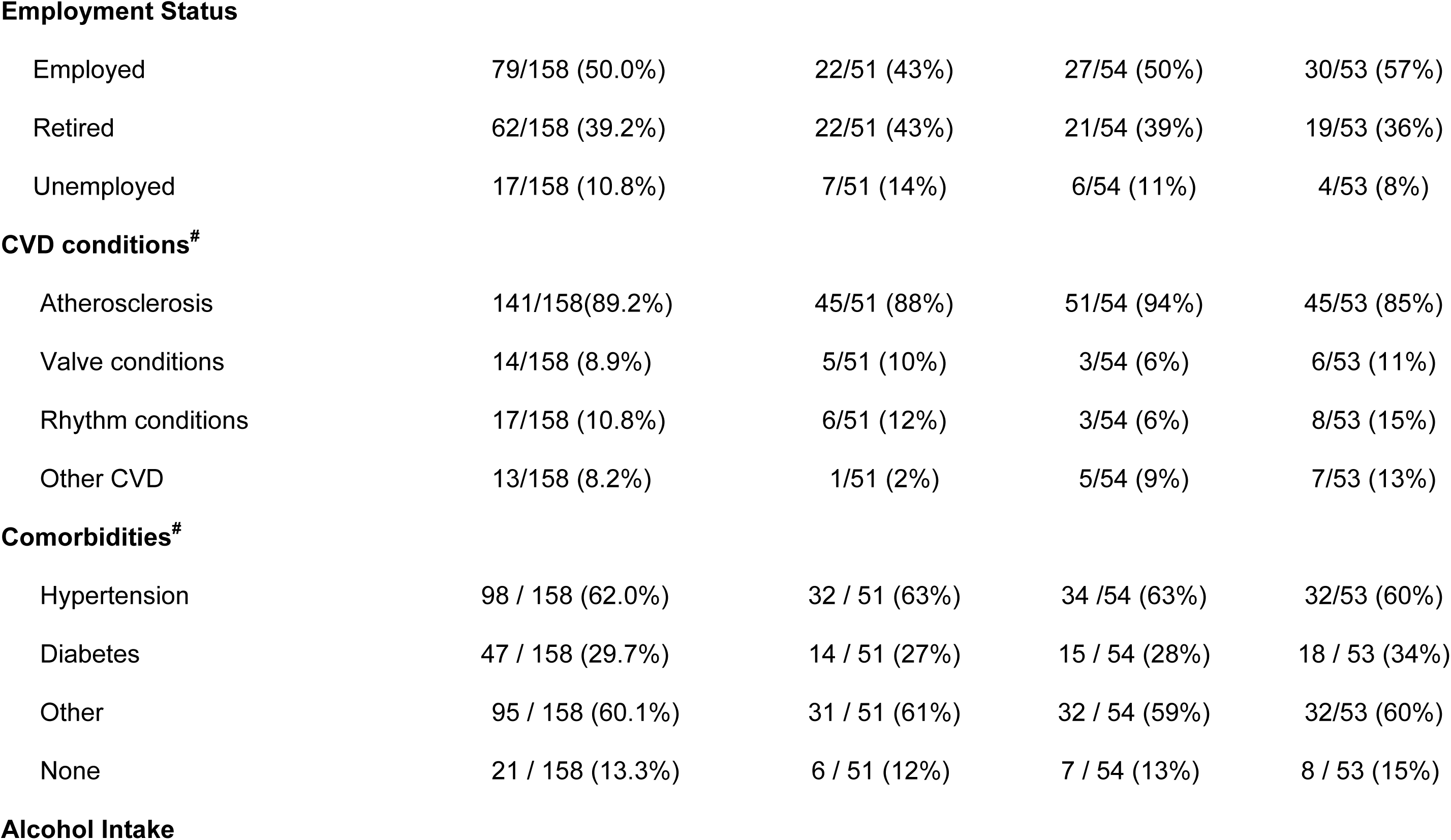
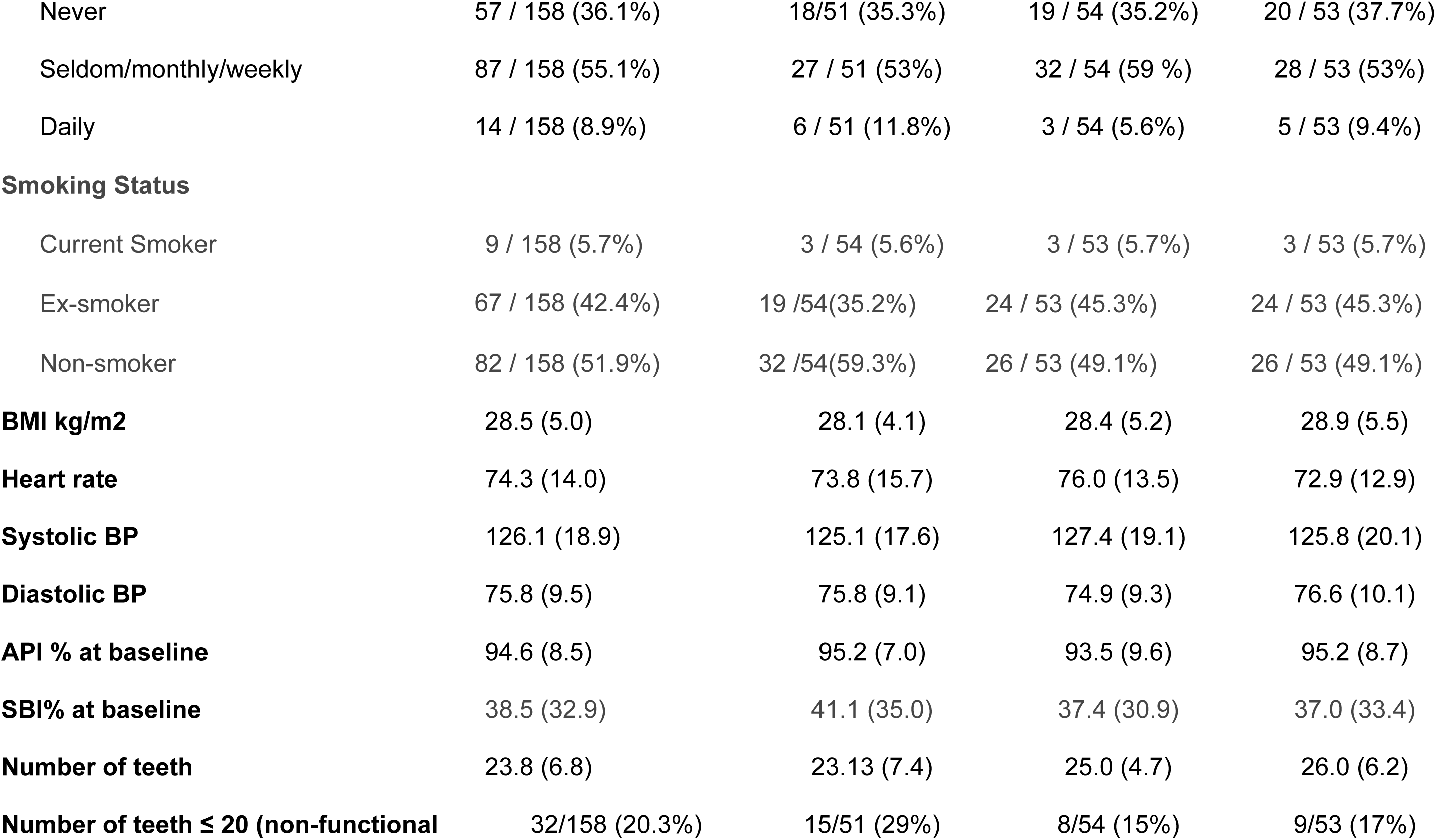
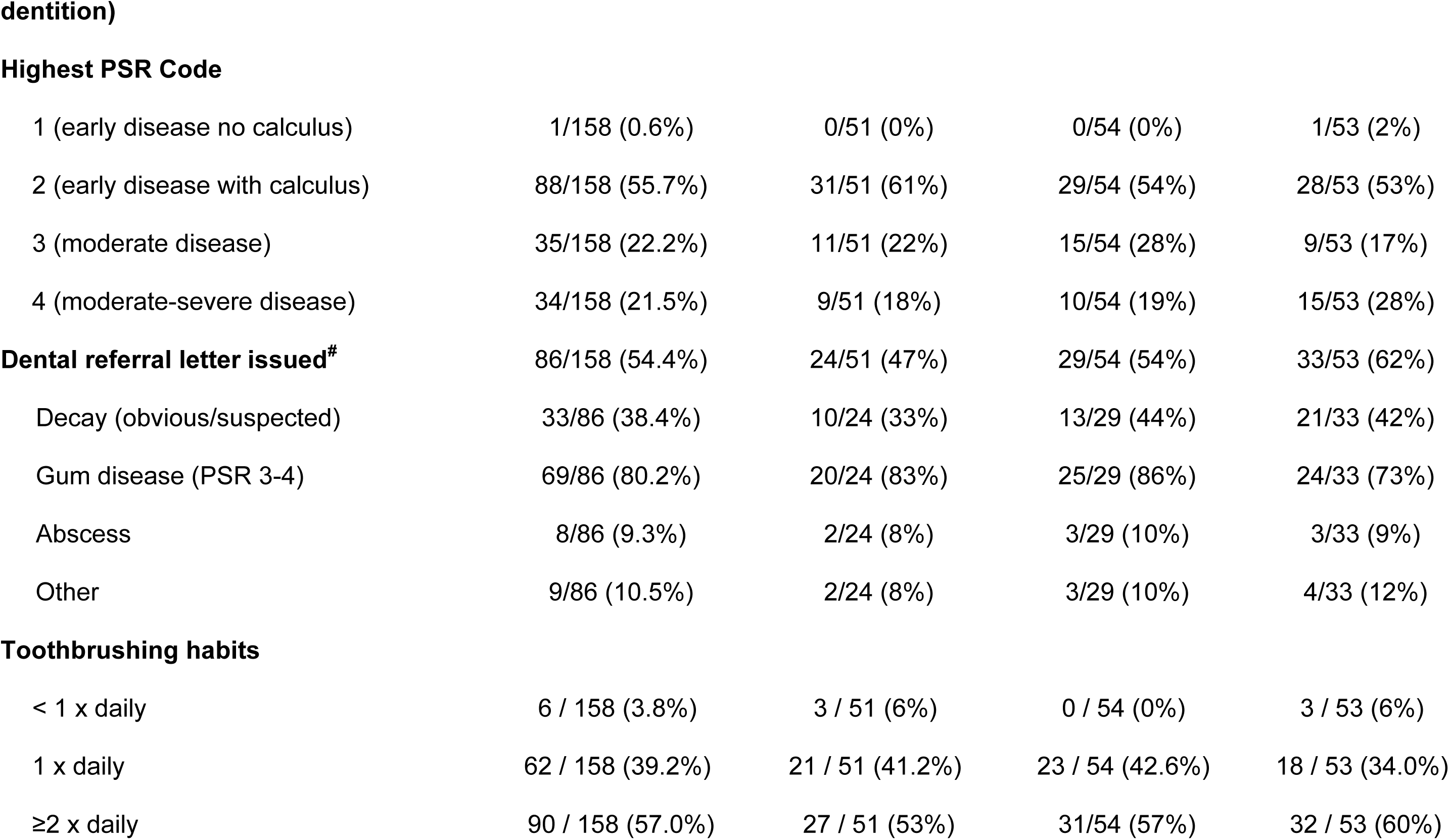
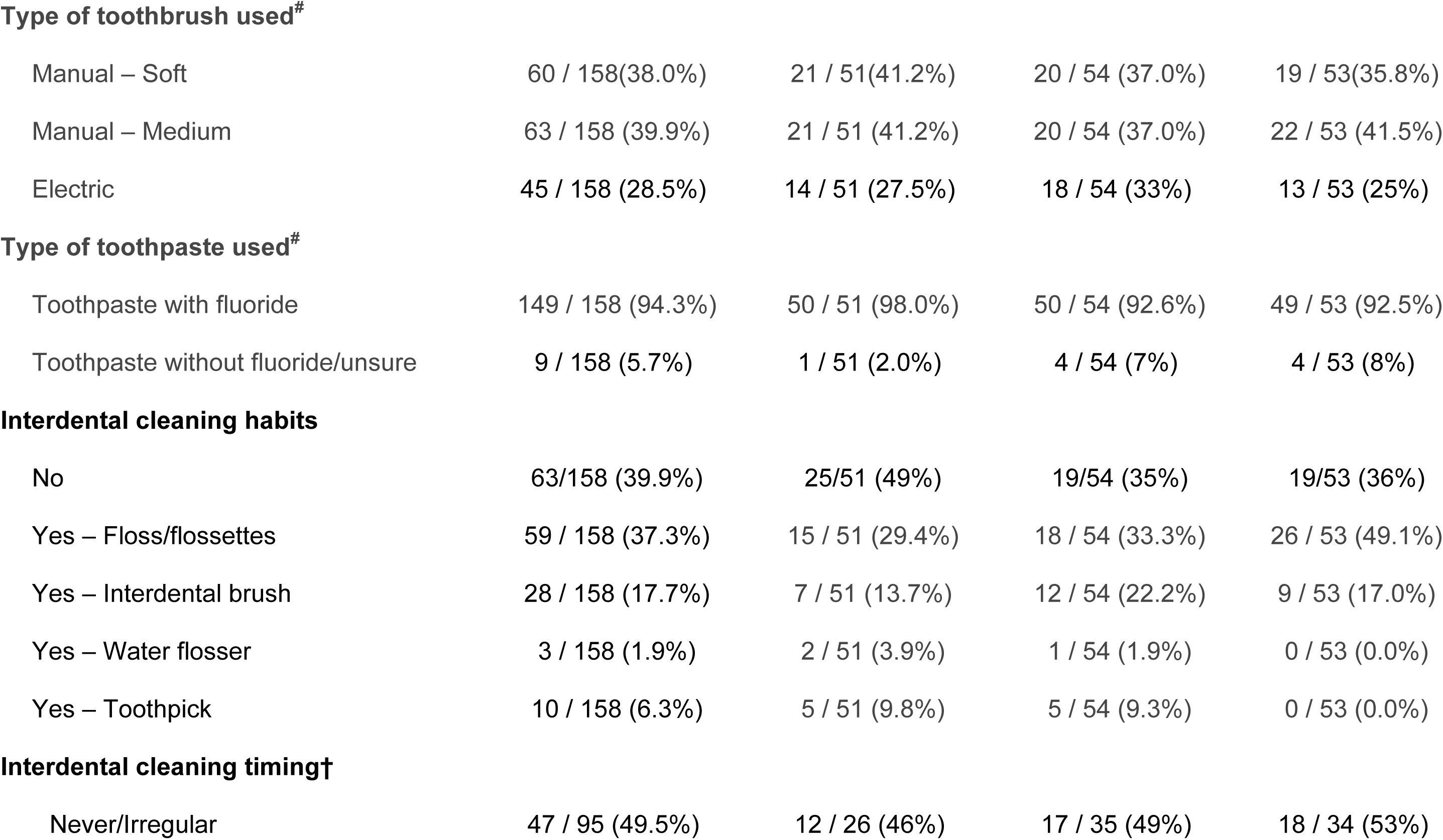
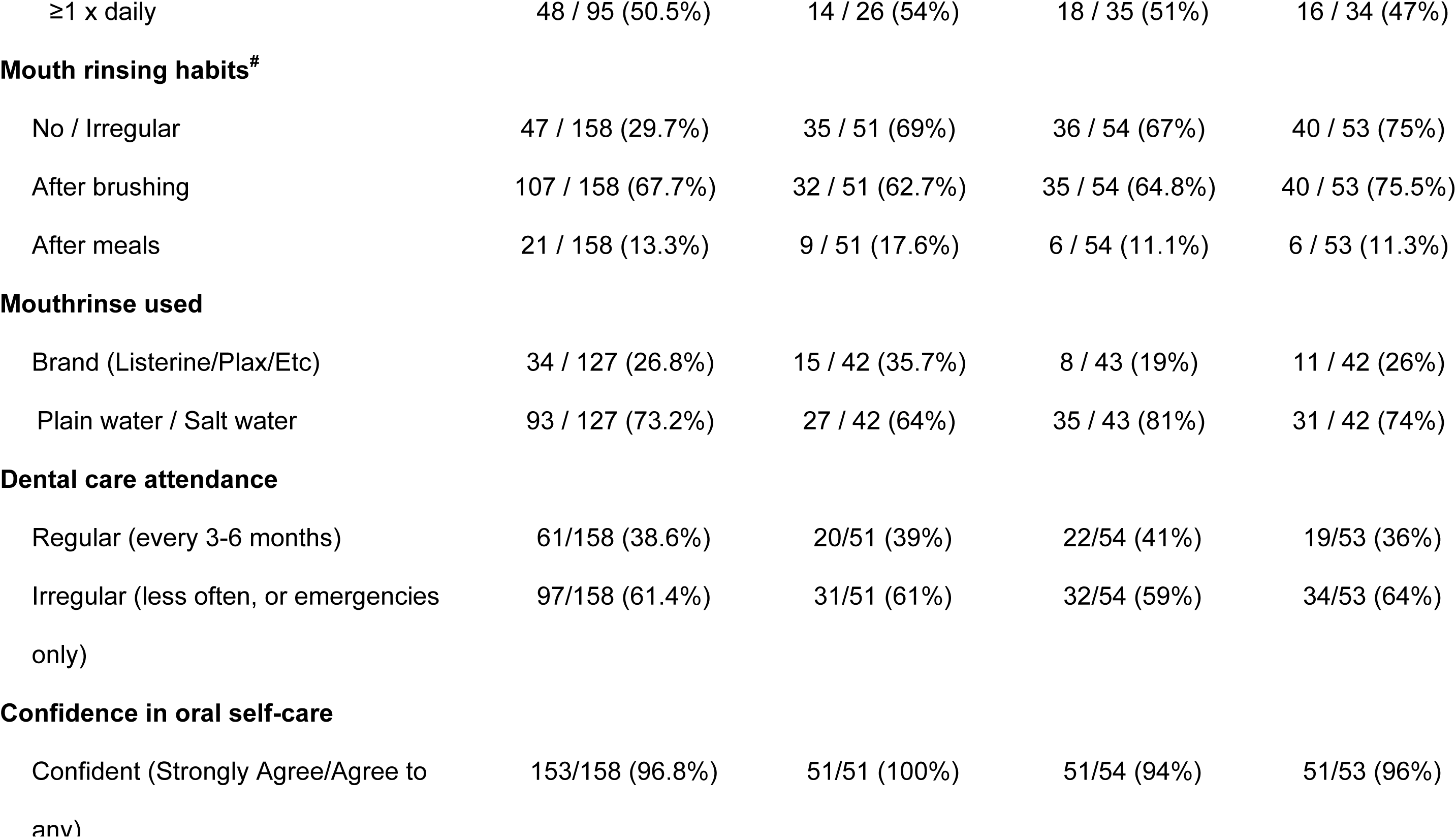
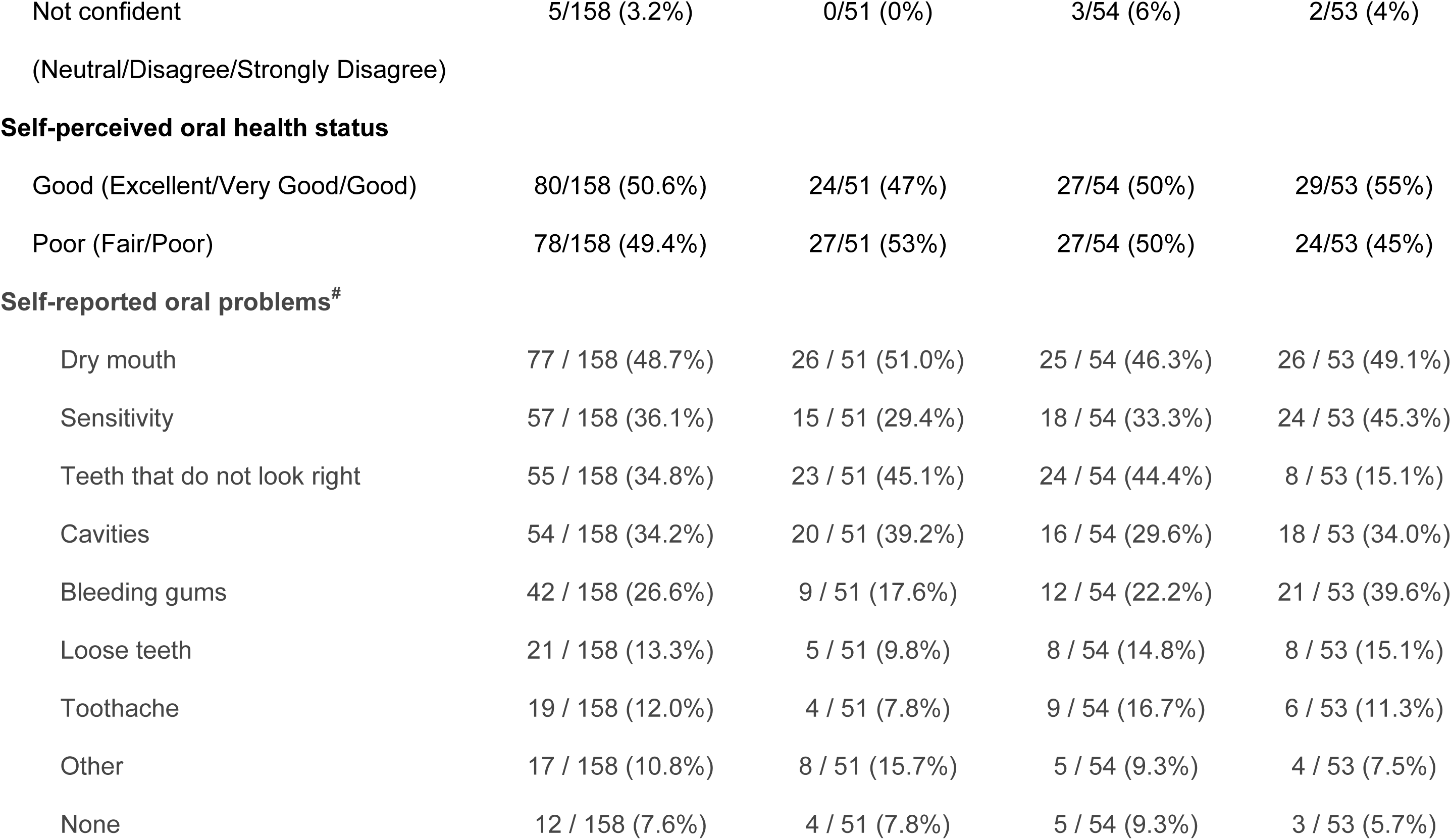
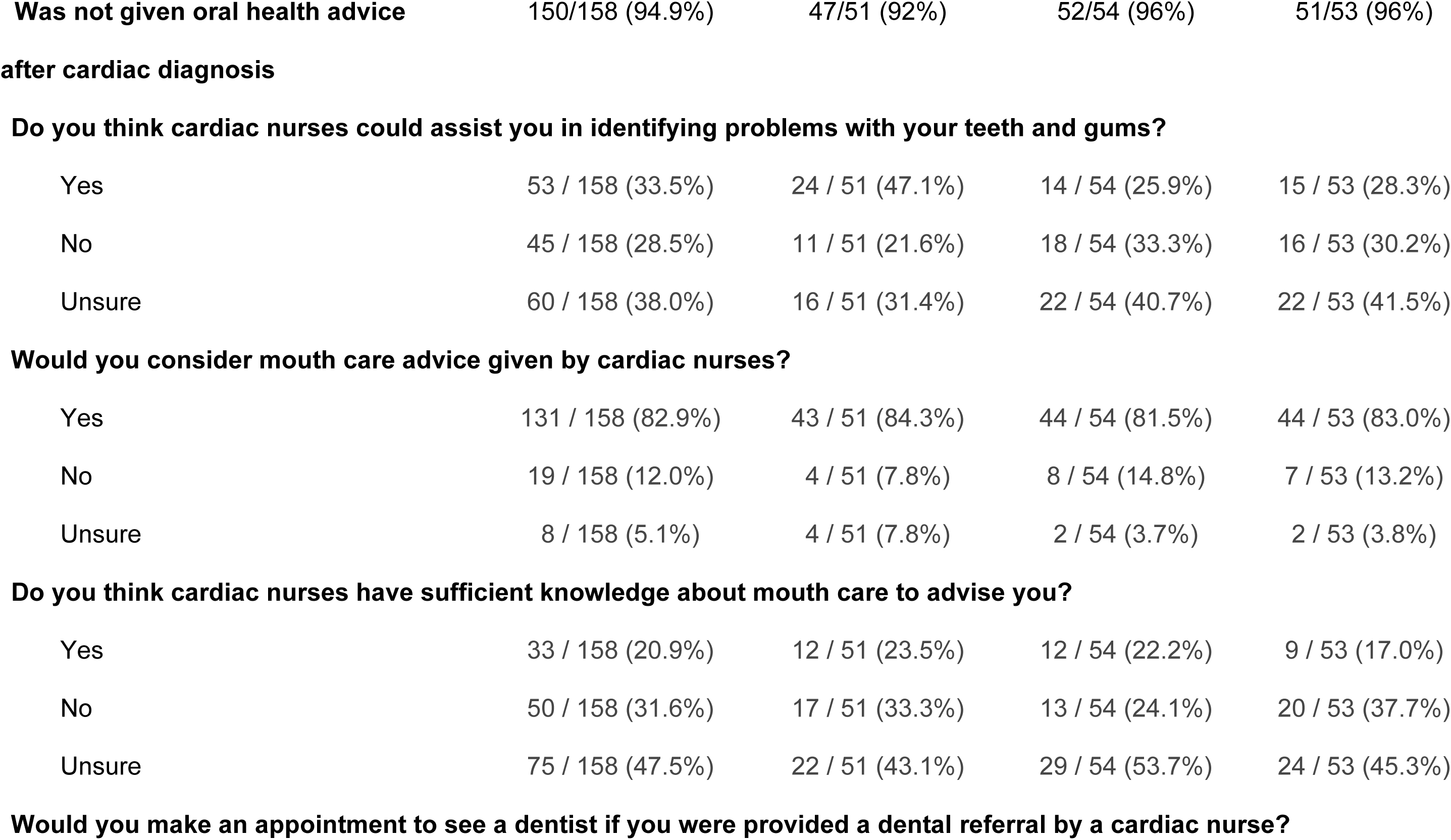
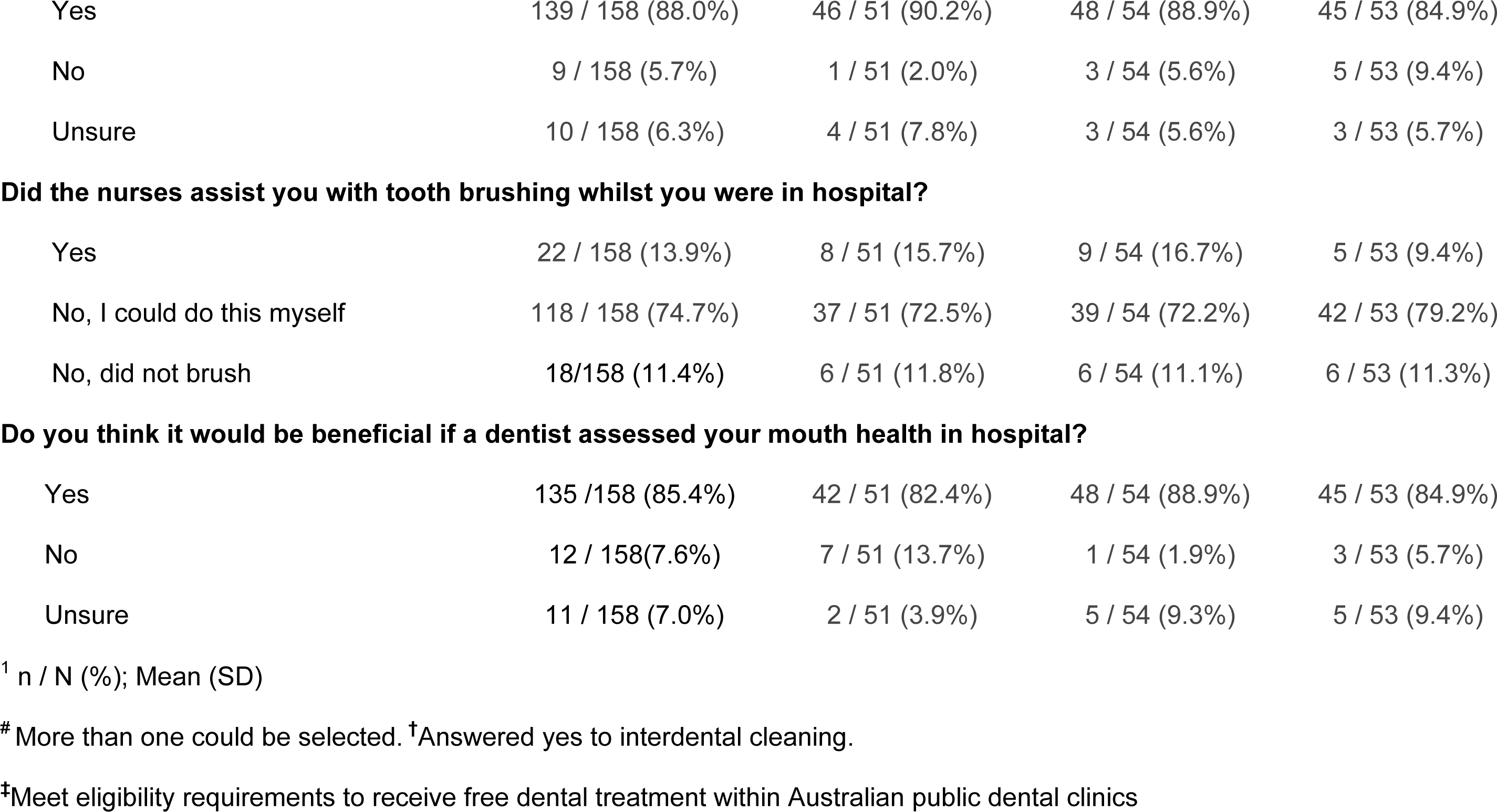
Baseline characteristics.

| **Characteristic** | **Overall**<br>N = 158<sup>1</sup> | **Control**<br>N = 51<sup>1</sup> | **OHI + DOHE**<br>N = 54<sup>1</sup> | **DOHE alone**<br>N = 53<sup>1</sup> |
| --- | --- | --- | --- | --- |
| **Location** |  |  |  |  |
| Westmead CEAP | 89/158 (56.3%) | 25/51 (49%) | 32/54 (59%) | 32/53 (60%) |
| Blacktown Mt Druitt CEAP | 69/158 (43.7%) | 26/51 (51%) | 22/54 (41%) | 21/53 (40%) |
| **Sex** |  |  |  |  |
| Male | 130/158 (82.3%) | 43/51 (84%) | 45/54 (83%) | 42/53 (79%) |
| **Age in years** | 62.0 (11.4) | 62.4 (11.6) | 63.3 (11.3) | 60.5 (11.5) |
| **Age Group** |  |  |  |  |
| 64 and under | 76/158 (48.1%) | 24/51 (47%) | 22/54 (41%) | 30/53 (57%) |
| 65 and over | 82/158 (51.9%) | 27/51 (53%) | 32/54 (59%) | 23/53 (43%) |
| **Ethnicity** |  |  |  |  |
| Asia (South, East, South-East) | 62/158 (39.2%) | 23/51 (45%) | 20/54 (37%) | 19/53 (36%) |
| Australia/New Zealand | 49/158 (31.0%) | 18/51 (35%) | 14/54 (26%) | 17/53 (32%) |
| Aboriginal and/or Torres Strait Islander | 3/158 (1.9%) | 1/51 (2%) | 1/54 (2%) | 1/53 (2%) |
| Other | 44/158 (27.8%) | 9/51 (18%) | 19/54 (35%) | 16/53 (30%) |
| **Income** |  |  |  |  |
| ≤\$49,000 | 49/158 (31.0%) | 17/51 (33%) | 16/54 (30%) | 16/53 (30%) |
| \$49,001 — \$80,000 | 23/158 (14.6%) | 11/51 (22%) | 8/54 (15%) | 4/53 (8%) |
| \$80,001 — \$120,000 | 18/158 (11.4%) | 6/51 (12%) | 6/54 (11%) | 6/53 (11%) |
| ≥\$120,001 | 34/158 (21.5%) | 10/51 (20%) | 9/54 (17%) | 15/53 (28%) |
| Prefer not to say | 34/158 (21.5%) | 7/51 (14%) | 15/54 (28%) | 12/53 (23%) |
| **Highest Education Qualification** |  |  |  |  |
| Up to primary school | 3/158 (1.9%) | 1/51 (2%) | 0/54 (0%) | 2/53 (4%) |
| Secondary school | 48/158 (30.4%) | 16/51 (31%) | 14/54 (26%) | 18/53 (34%) |
| Tertiary education | 107/158 (67.7%) | 34/51 (67%) | 40/54 (74%) | 33/53 (62%) |
| **Private Health** | 78/158 (49.4%) | 22/51 (43%) | 30/54 (56%) | 26/53 (49%) |
| **Eligible for Public Dentistry<sup>‡</sup>** |  |  |  |  |
| Eligible | 45/158 (28.5%) | 16/51 (31%) | 12/54 (22%) | 17/53 (32%) |
| Not eligible | 113/158 (71.5%) | 35/51 (69%) | 42/54 (78%) | 36/53 (68%) |
| **Employment Status** |  |  |  |  |
| Employed | 79/158 (50.0%) | 22/51 (43%) | 27/54 (50%) | 30/53 (57%) |
| Retired | 62/158 (39.2%) | 22/51 (43%) | 21/54 (39%) | 19/53 (36%) |
| Unemployed | 17/158 (10.8%) | 7/51 (14%) | 6/54 (11%) | 4/53 (8%) |
| **CVD conditions<sup>#</sup>** |  |  |  |  |
| Atherosclerosis | 141/158(89.2%) | 45/51 (88%) | 51/54 (94%) | 45/53 (85%) |
| Valve conditions | 14/158 (8.9%) | 5/51 (10%) | 3/54 (6%) | 6/53 (11%) |
| Rhythm conditions | 17/158 (10.8%) | 6/51 (12%) | 3/54 (6%) | 8/53 (15%) |
| Other CVD | 13/158 (8.2%) | 1/51 (2%) | 5/54 (9%) | 7/53 (13%) |
| **Comorbidities<sup>#</sup>** |  |  |  |  |
| Hypertension | 98 / 158 (62.0%) | 32 / 51 (63%) | 34 / 54 (63%) | 32/53 (60%) |
| Diabetes | 47 / 158 (29.7%) | 14 / 51 (27%) | 15 / 54 (28%) | 18 / 53 (34%) |
| Other | 95 / 158 (60.1%) | 31 / 51 (61%) | 32 / 54 (59%) | 32/53 (60%) |
| None | 21 / 158 (13.3%) | 6 / 51 (12%) | 7 / 54 (13%) | 8 / 53 (15%) |
| **Alcohol Intake** |  |  |  |  |
| Never | 57 / 158 (36.1%) | 18/51 (35.3%) | 19 / 54 (35.2%) | 20 / 53 (37.7%) |
| Seldom/monthly/weekly | 87 / 158 (55.1%) | 27 / 51 (53%) | 32 / 54 (59 %) | 28 / 53 (53%) |
| Daily | 14 / 158 (8.9%) | 6 / 51 (11.8%) | 3 / 54 (5.6%) | 5 / 53 (9.4%) |
| **Smoking Status** |  |  |  |  |
| Current Smoker | 9 / 158 (5.7%) | 3 / 54 (5.6%) | 3 / 53 (5.7%) | 3 / 53 (5.7%) |
| Ex-smoker | 67 / 158 (42.4%) | 19 /54(35.2%) | 24 / 53 (45.3%) | 24 / 53 (45.3%) |
| Non-smoker | 82 / 158 (51.9%) | 32 /54(59.3%) | 26 / 53 (49.1%) | 26 / 53 (49.1%) |
| **BMI kg/m2** | 28.5 (5.0) | 28.1 (4.1) | 28.4 (5.2) | 28.9 (5.5) |
| **Heart rate** | 74.3 (14.0) | 73.8 (15.7) | 76.0 (13.5) | 72.9 (12.9) |
| **Systolic BP** | 126.1 (18.9) | 125.1 (17.6) | 127.4 (19.1) | 125.8 (20.1) |
| **Diastolic BP** | 75.8 (9.5) | 75.8 (9.1) | 74.9 (9.3) | 76.6 (10.1) |
| **API % at baseline** | 94.6 (8.5) | 95.2 (7.0) | 93.5 (9.6) | 95.2 (8.7) |
| **SBI% at baseline** | 38.5 (32.9) | 41.1 (35.0) | 37.4 (30.9) | 37.0 (33.4) |
| **Number of teeth** | 23.8 (6.8) | 23.13 (7.4) | 25.0 (4.7) | 26.0 (6.2) |
| **Number of teeth ≤ 20 (non-functional** | 32/158 (20.3%) | 15/51 (29%) | 8/54 (15%) | 9/53 (17%) |
| **Highest PSR Code** |  |  |  |  |
| 1 (early disease no calculus) | 1/158 (0.6%) | 0/51 (0%) | 0/54 (0%) | 1/53 (2%) |
| 2 (early disease with calculus) | 88/158 (55.7%) | 31/51 (61%) | 29/54 (54%) | 28/53 (53%) |
| 3 (moderate disease) | 35/158 (22.2%) | 11/51 (22%) | 15/54 (28%) | 9/53 (17%) |
| 4 (moderate-severe disease) | 34/158 (21.5%) | 9/51 (18%) | 10/54 (19%) | 15/53 (28%) |
| **Dental referral letter issued<sup>#</sup>** |  |  |  |  |
| Decay (obvious/suspected) | 33/86 (38.4%) | 10/24 (33%) | 13/29 (44%) | 21/33 (42%) |
| Gum disease (PSR 3-4) | 69/86 (80.2%) | 20/24 (83%) | 25/29 (86%) | 24/33 (73%) |
| Abscess | 8/86 (9.3%) | 2/24 (8%) | 3/29 (10%) | 3/33 (9%) |
| Other | 9/86 (10.5%) | 2/24 (8%) | 3/29 (10%) | 4/33 (12%) |
| **Toothbrushing habits** |  |  |  |  |
| < 1 x daily | 6 / 158 (3.8%) | 3 / 51 (6%) | 0 / 54 (0%) | 3 / 53 (6%) |
| 1 x daily | 62 / 158 (39.2%) | 21 / 51 (41.2%) | 23 / 54 (42.6%) | 18 / 53 (34.0%) |
| ≥2 x daily | 90 / 158 (57.0%) | 27 / 51 (53%) | 31/54 (57%) | 32 / 53 (60%) |
| **Type of toothbrush used<sup>#</sup>** |  |  |  |  |
| Manual – Soft | 60 / 158(38.0%) | 21 / 51(41.2%) | 20 / 54 (37.0%) | 19 / 53(35.8%) |
| Manual – Medium | 63 / 158 (39.9%) | 21 / 51 (41.2%) | 20 / 54 (37.0%) | 22 / 53 (41.5%) |
| Electric | 45 / 158 (28.5%) | 14 / 51 (27.5%) | 18 / 54 (33%) | 13 / 53 (25%) |
| **Type of toothpaste used<sup>#</sup>** |  |  |  |  |
| Toothpaste with fluoride | 149 / 158 (94.3%) | 50 / 51 (98.0%) | 50 / 54 (92.6%) | 49 / 53 (92.5%) |
| Toothpaste without fluoride/unsure | 9 / 158 (5.7%) | 1 / 51 (2.0%) | 4 / 54 (7%) | 4 / 53 (8%) |
| **Interdental cleaning habits** |  |  |  |  |
| No | 63/158 (39.9%) | 25/51 (49%) | 19/54 (35%) | 19/53 (36%) |
| Yes – Floss/flossettes | 59 / 158 (37.3%) | 15 / 51 (29.4%) | 18 / 54 (33.3%) | 26 / 53 (49.1%) |
| Yes – Interdental brush | 28 / 158 (17.7%) | 7 / 51 (13.7%) | 12 / 54 (22.2%) | 9 / 53 (17.0%) |
| Yes – Water flosser | 3 / 158 (1.9%) | 2 / 51 (3.9%) | 1 / 54 (1.9%) | 0 / 53 (0.0%) |
| Yes – Toothpick | 10 / 158 (6.3%) | 5 / 51 (9.8%) | 5 / 54 (9.3%) | 0 / 53 (0.0%) |
| **Interdental cleaning timing†** |  |  |  |  |
| Never/Irregular | 47 / 95 (49.5%) | 12 / 26 (46%) | 17 / 35 (49%) | 18 / 34 (53%) |
| ≥1 x daily | 48 / 95 (50.5%) | 14 / 26 (54%) | 18 / 35 (51%) | 16 / 34 (47%) |
| **Mouth rinsing habits<sup>#</sup>** |  |  |  |  |
| No / Irregular | 47 / 158 (29.7%) | 35 / 51 (69%) | 36 / 54 (67%) | 40 / 53 (75%) |
| After brushing | 107 / 158 (67.7%) | 32 / 51 (62.7%) | 35 / 54 (64.8%) | 40 / 53 (75.5%) |
| After meals | 21 / 158 (13.3%) | 9 / 51 (17.6%) | 6 / 54 (11.1%) | 6 / 53 (11.3%) |
| **Mouthrinse used** |  |  |  |  |
| Brand (Listerine/Plax/Etc) | 34 / 127 (26.8%) | 15 / 42 (35.7%) | 8 / 43 (19%) | 11 / 42 (26%) |
| Plain water / Salt water | 93 / 127 (73.2%) | 27 / 42 (64%) | 35 / 43 (81%) | 31 / 42 (74%) |
| **Dental care attendance** |  |  |  |  |
| Regular (every 3-6 months) | 61/158 (38.6%) | 20/51 (39%) | 22/54 (41%) | 19/53 (36%) |
| Irregular (less often, or emergencies only) | 97/158 (61.4%) | 31/51 (61%) | 32/54 (59%) | 34/53 (64%) |
| **Confidence in oral self-care** |  |  |  |  |
| Confident (Strongly Agree/Agree to any) | 153/158 (96.8%) | 51/51 (100%) | 51/54 (94%) | 51/53 (96%) |
| Not confident | 5/158 (3.2%) | 0/51 (0%) | 3/54 (6%) | 2/53 (4%) |
| (Neutral/Disagree/Strongly Disagree) |  |  |  |  |
| **Self-perceived oral health status** |  |  |  |  |
| Good (Excellent/Very Good/Good) | 80/158 (50.6%) | 24/51 (47%) | 27/54 (50%) | 29/53 (55%) |
| Poor (Fair/Poor) | 78/158 (49.4%) | 27/51 (53%) | 27/54 (50%) | 24/53 (45%) |
| **Self-reported oral problems<sup>#</sup>** |  |  |  |  |
| Dry mouth | 77 / 158 (48.7%) | 26 / 51 (51.0%) | 25 / 54 (46.3%) | 26 / 53 (49.1%) |
| Sensitivity | 57 / 158 (36.1%) | 15 / 51 (29.4%) | 18 / 54 (33.3%) | 24 / 53 (45.3%) |
| Teeth that do not look right | 55 / 158 (34.8%) | 23 / 51 (45.1%) | 24 / 54 (44.4%) | 8 / 53 (15.1%) |
| Cavities | 54 / 158 (34.2%) | 20 / 51 (39.2%) | 16 / 54 (29.6%) | 18 / 53 (34.0%) |
| Bleeding gums | 42 / 158 (26.6%) | 9 / 51 (17.6%) | 12 / 54 (22.2%) | 21 / 53 (39.6%) |
| Loose teeth | 21 / 158 (13.3%) | 5 / 51 (9.8%) | 8 / 54 (14.8%) | 8 / 53 (15.1%) |
| Toothache | 19 / 158 (12.0%) | 4 / 51 (7.8%) | 9 / 54 (16.7%) | 6 / 53 (11.3%) |
| Other | 17 / 158 (10.8%) | 8 / 51 (15.7%) | 5 / 54 (9.3%) | 4 / 53 (7.5%) |
| None | 12 / 158 (7.6%) | 4 / 51 (7.8%) | 5 / 54 (9.3%) | 3 / 53 (5.7%) |
| **Was not given oral health advice** | 150/158 (94.9%) | 47/51 (92%) | 52/54 (96%) | 51/53 (96%) |
| **after cardiac diagnosis** |  |  |  |  |
| **Do you think cardiac nurses could assist you in identifying problems with your teeth and gums?** |  |  |  |  |
| Yes | 53 / 158 (33.5%) | 24 / 51 (47.1%) | 14 / 54 (25.9%) | 15 / 53 (28.3%) |
| No | 45 / 158 (28.5%) | 11 / 51 (21.6%) | 18 / 54 (33.3%) | 16 / 53 (30.2%) |
| Unsure | 60 / 158 (38.0%) | 16 / 51 (31.4%) | 22 / 54 (40.7%) | 22 / 53 (41.5%) |
| **Would you consider mouth care advice given by cardiac nurses?** |  |  |  |  |
| Yes | 131 / 158 (82.9%) | 43 / 51 (84.3%) | 44 / 54 (81.5%) | 44 / 53 (83.0%) |
| No | 19 / 158 (12.0%) | 4 / 51 (7.8%) | 8 / 54 (14.8%) | 7 / 53 (13.2%) |
| Unsure | 8 / 158 (5.1%) | 4 / 51 (7.8%) | 2 / 54 (3.7%) | 2 / 53 (3.8%) |
| **Do you think cardiac nurses have sufficient knowledge about mouth care to advise you?** |  |  |  |  |
| Yes | 33 / 158 (20.9%) | 12 / 51 (23.5%) | 12 / 54 (22.2%) | 9 / 53 (17.0%) |
| No | 50 / 158 (31.6%) | 17 / 51 (33.3%) | 13 / 54 (24.1%) | 20 / 53 (37.7%) |
| Unsure | 75 / 158 (47.5%) | 22 / 51 (43.1%) | 29 / 54 (53.7%) | 24 / 53 (45.3%) |
| **Would you make an appointment to see a dentist if you were provided a dental referral by a cardiac nurse?** |  |  |  |  |
| Yes | 139 / 158 (88.0%) | 46 / 51 (90.2%) | 48 / 54 (88.9%) | 45 / 53 (84.9%) |
| No | 9 / 158 (5.7%) | 1 / 51 (2.0%) | 3 / 54 (5.6%) | 5 / 53 (9.4%) |
| Unsure | 10 / 158 (6.3%) | 4 / 51 (7.8%) | 3 / 54 (5.6%) | 3 / 53 (5.7%) |
| **Did the nurses assist you with tooth brushing whilst you were in hospital?** |  |  |  |  |
| Yes | 22 / 158 (13.9%) | 8 / 51 (15.7%) | 9 / 54 (16.7%) | 5 / 53 (9.4%) |
| No, I could do this myself | 118 / 158 (74.7%) | 37 / 51 (72.5%) | 39 / 54 (72.2%) | 42 / 53 (79.2%) |
| No, did not brush | 18/158 (11.4%) | 6 / 51 (11.8%) | 6 / 54 (11.1%) | 6 / 53 (11.3%) |
| **Do you think it would be beneficial if a dentist assessed your mouth health in hospital?** |  |  |  |  |
| Yes | 135 /158 (85.4%) | 42 / 51 (82.4%) | 48 / 54 (88.9%) | 45 / 53 (84.9%) |
| No | 12 / 158(7.6%) | 7 / 51 (13.7%) | 1 / 54 (1.9%) | 3 / 53 (5.7%) |
| Unsure | 11 / 158 (7.0%) | 2 / 51 (3.9%) | 5 / 54 (9.3%) | 5 / 53 (9.4%) |

<sup>1</sup> n / N (%); Mean (SD)
<sup>#</sup> More than one could be selected. <sup>†</sup>Answered yes to interdental cleaning.
<sup>‡</sup>Meet eligibility requirements to receive free dental treatment within Australian public dental clinics

At baseline, 57% of participants brushed their teeth twice daily or more, 50.5% cleaned between their teeth once daily and over 70% used a manual toothbrush. The majority, 70.1% rinsed with mouthwash or water immediately after brushing. The mean API was 94.6±8.5, and the mean SBI 38.5±32.9 percent.

Regarding oral disease burden, 20.3% had fewer than twenty teeth, 43.6% required treatment for moderate to severe periodontal disease and 48% were experiencing dry mouth. Thirty-eight percent reported regular dental service use, and referral letters for dental conditions (dental decay, abscesses, or other concerns) identified at baseline were issued to 54.4% of participants.

### Impact of oral health education on oral hygiene

At 6-weeks 77.1% (27/35) of participants receiving OHI+DOHE reduced their API from baseline, compared to 26.8% (11/51) in the usual care group (OR, 9.16 [95%CI:3.34-27.75] p<0.001) (Table 2). The primary outcome was missing for 25% of participants which was more common for those with private health insurance and among those identifying as Asian heritage (Table S4 Supplementary File 5). However, sensitivity analyses found a consistent result (worst-case method (OR, 3.59 [95%CI:1.55-8.74] p=0.004), MICE method (OR, 6.66 [95%CI: 2.33-19.07] p<0.001) (Table S5 Supplementary File 5)). Follow-up was low at 12 weeks and found only those with the OHI+DOHE group maintained a significantly reduced API score.

**Table 2.** Reduced API and changes to oral health behaviours from baseline.

|  | **Control**<br>**N = 51** | **OHI+DOHE**<br>**N = 54** | **DOHE alone**<br>**N = 53** | **OR**<br>**(95%CI)** | **p-value** |
| --- | --- | --- | --- | --- | --- |
| **Reduced API by any amount from baseline level** |  |  |  |  |  |
| **6 weeks** | 26.8% (11/41) | 77.1% (27/35) | 63.4% (26/41) | **9.16 (3.3-7.7)<sup>1\*</sup>** | <0.001 |
|  | (14.8%-43.2%) | (59.4%-9.0%) | (46.9%-7.4%) | 4.72 (1.8-12.5)<sup>2</sup> | <0.001 |
|  |  |  |  | 0.49 (0.1-1.3)<sup>3</sup> | 0.2 |
| **12 weeks** | 27.8% (5/18) | 83.3% (15/18) | 47.4% (9/19) | 17.2 (3.3-144)<sup>1</sup> | <0.002 |
|  | (10.7%-53.6%) | (57.7%-5.6%) | (25.2%-70.5%) | 1.95 (0.4-8.9)<sup>2</sup> | 0.4 |
|  |  |  |  | 0.69 (0.2-2.3)<sup>3</sup> | 0.6 |
| **Have you changed any mouth care habits?** |  |  |  |  |  |
| **6-Weeks** | 17.8% (8/45) | 70.5% (31/44) | 55.8% (24/43) | 11.74 (4.3-35.8)<sup>1</sup> | <0.001 |
|  | (8.5%-32.6%) | (54.6%- 82.8%) | (40.0%-70.6%) | 6.23 (2.3-18.2)<sup>2</sup> | <0.001 |
|  |  |  |  | 1.89 (0.8-4.9)<sup>3</sup> | 0.2 |
| **12-weeks** | 20.8% (5/24) | 58.3% (14/24) | 66.7% (16/24) | 6.16(1.7-25.8)<sup>1</sup> | 0.008 |
|  | (7.9%-42.7%) | (36.9%-77.2%) | (44.7%-83.6%) | 9.36(2.5-42.4)<sup>2</sup> | 0.002 |
|  |  |  |  | 0.69 (0.2-2.3)<sup>3</sup> | 0.6 |

\*Primary outcome, <sup>1</sup> Control versus OHI+DOHE, <sup>2</sup> Control versus DOHE alone, <sup>3</sup> OHI+DOHE versus DOHE alone

The secondary comparison of DOHE alone versus usual care at 6-weeks found that 63.4% (26/41) versus 26.8% (11/41) reduced their API (OR, 4.72 [95%CI: 1.8-12.5] p<0.001), although this attenuated at 12-weeks (Table 2). There was no evidence of a significant difference between OHI+DOHE or DOHE alone.

Mean API reduced and mean SBI increased in all groups (Table S6 Supplementary File 5). At 6 weeks, none of the participants in the control group and one participant from each intervention group achieved optimal oral hygiene (API score of ≤35%) (Table S7 Supplementary File 5). There was no difference between groups in participants who reached optimal gingival inflammation (SBI score of ≤ 25%) at either 6- or 12-weeks post baseline.

### Impact of oral health education on oral health behaviours and knowledge

At 6 weeks, oral hygiene behaviours improved in both intervention groups compared to the usual care group. A total of 70.5% (31/44) of participants receiving OHI+DOHE and 55.8% (24/43) of participants receiving DOHE alone reported changes to their oral hygiene behaviours, compared to 17.8% (8/45) of participants receiving usual care, (OR, 11.74 [95% CI:4.3-35.8] p<0.001) and (OR, 6.23 [95% CI:2.3-18.2] p<0.001) respectively (Table 2). These behavioural changes included changes in frequency, duration, and technique of toothbrushing and/or cleaning between teeth. This pattern continued for the subset proceeding to 12-weeks, see Table 2.

Knowledge regarding the impact oral health has on heart health improved across all study groups (Figure 2) with larger increases in the two intervention groups 61% (27/44) (95% CI: 46–75%) of those receiving OHI+DOHE, 77% (33/43) (95% CI: 61–88%) of those receiving DOHE alone compared to 41% (18/44) (95% CI: 27–57%) of those receiving usual care (p=0.003) (Table S8 Supplementary File 5). These differences did not remain significant in the subset that continued to 12-weeks. The question ‘poor mouth health may affect an existing heart condition’ showed the largest difference [90.9% (40/44) (95% CI: 77.4-97.0%) OHI+DOHE group, 93.0% (40/43) (95% CI:79.9-98.2%) DOHE alone group, and 61.4% (27/44) (95% CI: 45.5-75.3%) in the usual care group] (Table 9 Supplementary File 5).

**Figure 2.**
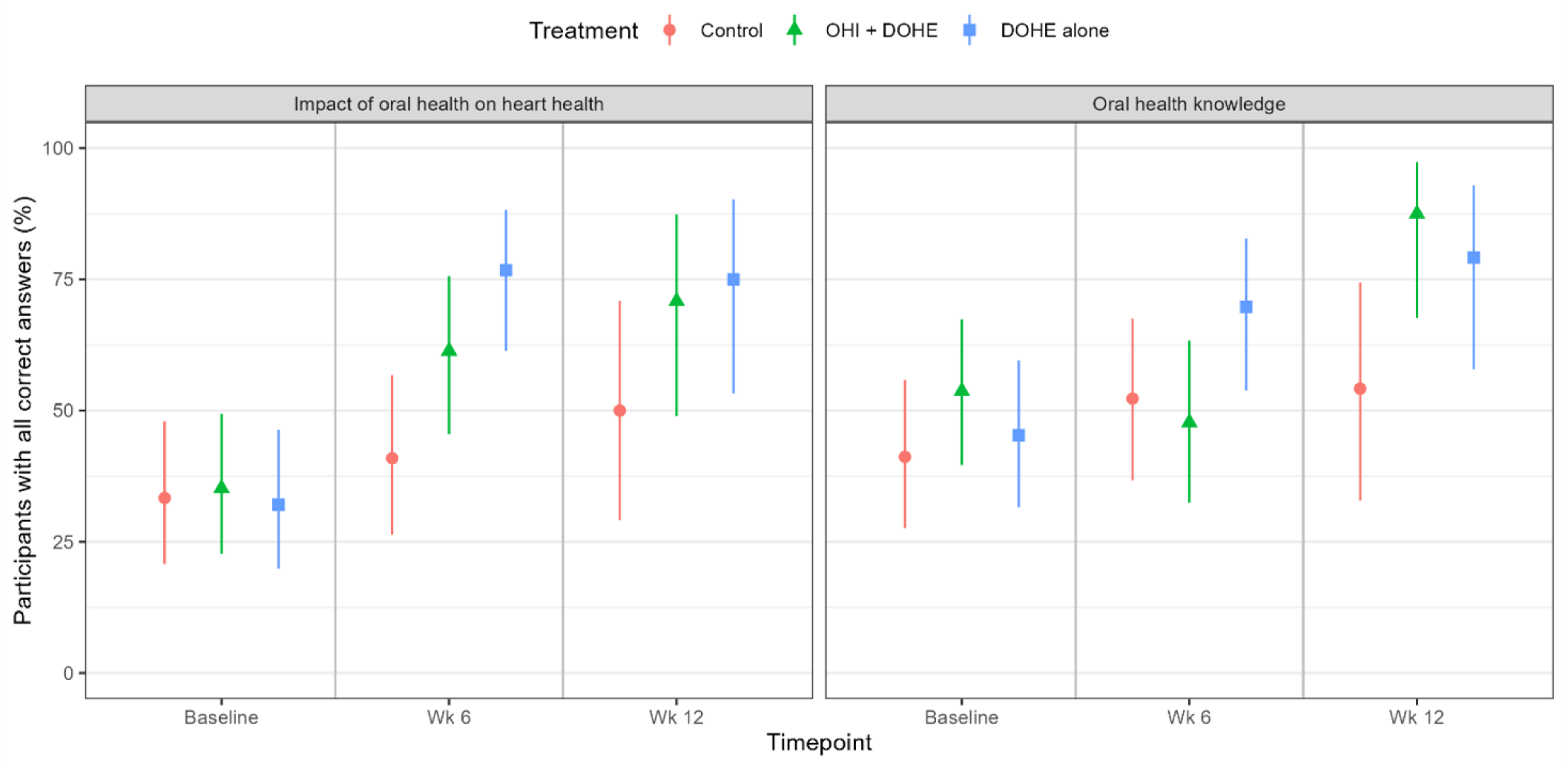
Correct answers to domains of the CVD Knowledge Questionnaire Knowledge regarding the impact oral health has on heart health improved.

Although general oral health knowledge improved across all study arms with 52% (23/44) (95% CI: 37–67%) of those receiving OHI+DOHE, 70% (30/43) (95% CI: 54–82%) of those receiving DOHE alone and 48% (21/44) (95% CI: 33–63%) of those receiving usual care scoring higher on these questions at 6 weeks, these changes were not significant (p=0.092) (Figure 2) Table S8 Supplementary File 5). They became significant in the subset that continued to 12-weeks with larger increases in both intervention groups compared to the usual care group. Knowledge regarding the oral implications of heart medication was low at baseline, with minimal improvement throughout the study period (Table S8 Supplementary File 5).

### Participant perceptions of oral health

The intervention did not impact confidence in oral self-care which was high at baseline in all study arms and was unchanged at follow-up (Table 3). Similarly, self-reported oral health status and motivation to make a dental appointment were not impacted by the intervention at either 6 or 12 weeks (Table 4). However, despite no influence from the interventions, 79% of the cohort were motivated to see an oral health practitioner at each follow-up [6-weeks 104/132 (95% CI:71-85%); 12-weeks 57/72 (95% CI: 68-88%)] (Table 5).

**Table 3.**
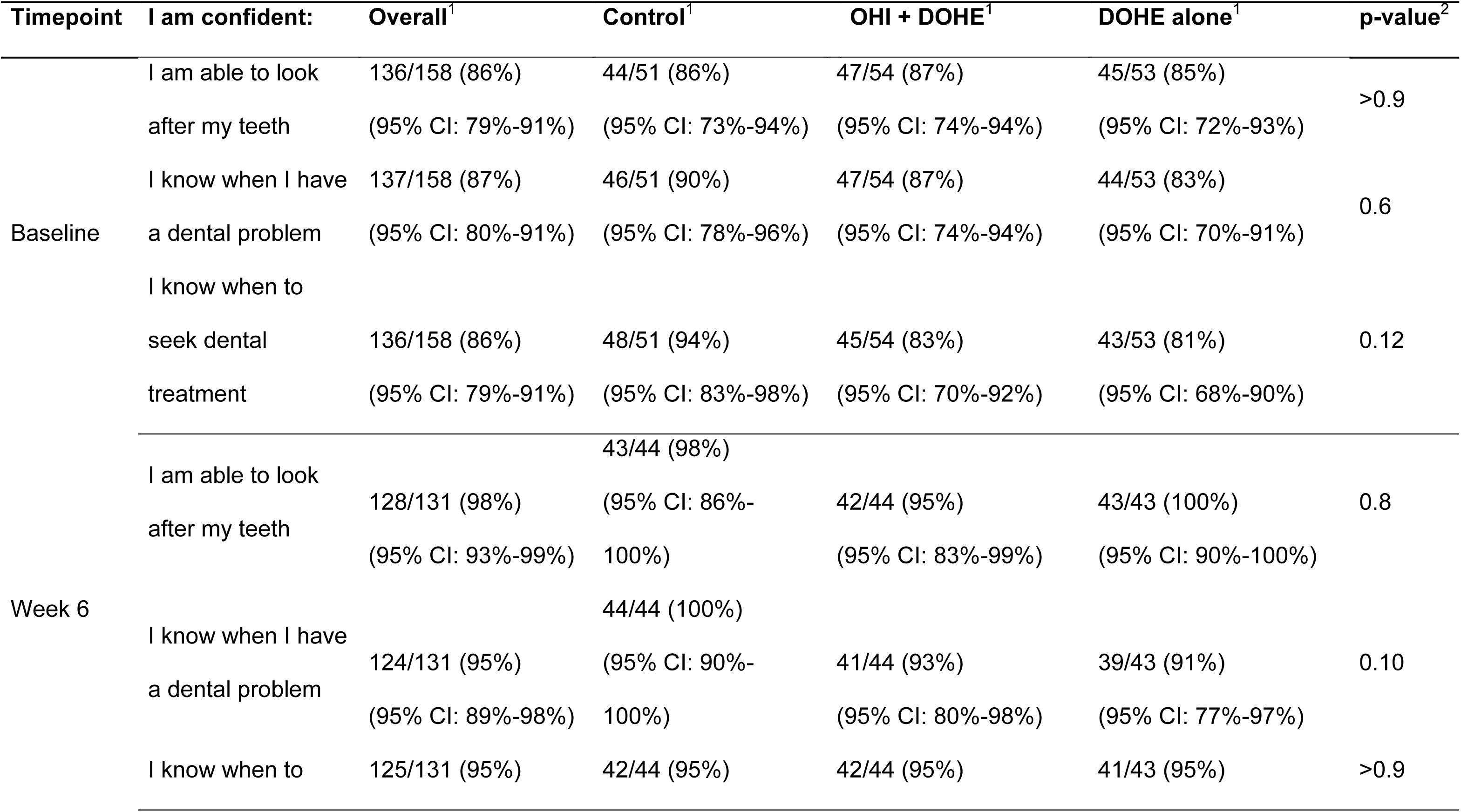
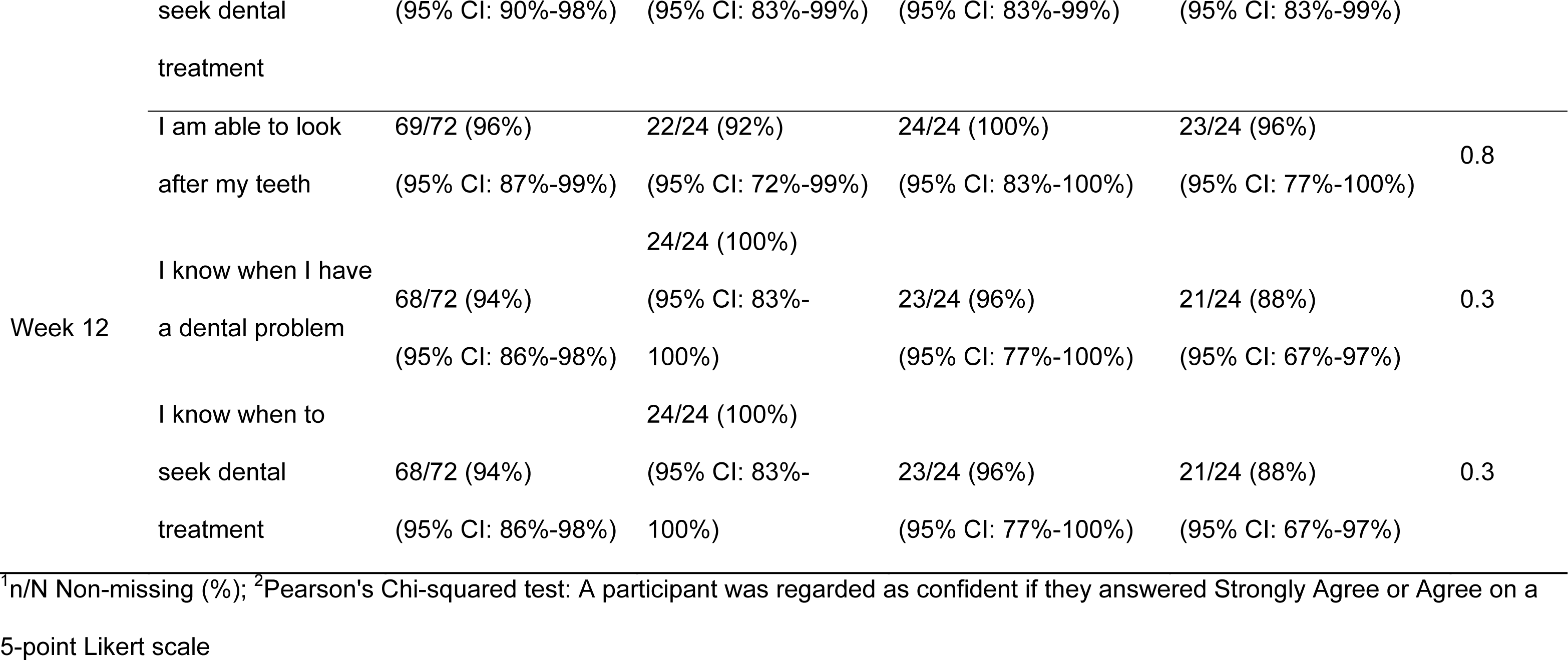
Confidence in oral self-care by timepoint.

**Table 4.** Changes in perceived oral health status by timepoint.

| Timepoint | Characteristic | Overall<sup>1</sup> | Control<sup>1</sup> | OHI+DOHE<sup>1</sup> | DOHE alone<sup>1</sup> | p-value<sup>2</sup> |
| --- | --- | --- | --- | --- | --- | --- |
| Baseline | Positive self-perceived oral health status | 80/158 (51%)<br>(43-59%) | 24/51 (47%)<br>(33-61%) | 27/54 (50%)<br>(37-63%) | 29/53 (55%)<br>(41-68%) | 0.7 |
| Week 6 | Positive self-perceived oral health status | 39/48 (81%)<br>(67- 91%) | 12/16 (75%)<br>(47-92%) | 16/18 (89%)<br>(64-98%) | 11/14 (79%)<br>(49-94%) | 0.6 |
| Week 12 | Positive self-perceived oral health status | 31/35 (89%)<br>(72-96%) | 7/8 (88%)<br>(47-99%) | 15/15 (100%)<br>(75-100%) | 9/12 (75%)<br>(43-93%) | 0.10 |

<sup>1</sup>n/N Non-missing (%), <sup>2</sup>Pearson's Chi-squared test. This table only the percentage of the study population that answered 'yes' to the question "Have you noticed an improvement of your gum health since last visit" and then went on to change their oral health status answer to good or higher at follow-up.

**Table 5.** Participants who were motivated to see a dental practitioner, by timepoint.

| Characteristic | Overall<sup>1</sup> | Control<sup>1</sup> | OHI + DOHE<sup>1</sup> | DOHE alone<sup>1</sup> | p-value<sup>2</sup> |
| --- | --- | --- | --- | --- | --- |
| Motivation 6-Weeks |  |  |  |  |  |
| I am motivated to visit a dentist regularly | 104/132 (79%)<br>(95% CI:71-85%) | 32/45 (71%)<br>(95% CI:55-83%) | 37/44 (84%)<br>(95% CI: 69-93%) | 35/43 (81%)<br>(95% CI: 66-91%) | 0.3 |
| Motivation 12-weeks |  |  |  |  |  |
| I am motivated to visit a dentist regularly | 57/72 (79%)<br>(95% CI: 68-88%) | 19/24 (79%)<br>(95% CI: 57-92%) | 19/24 (79%)<br>(95% CI: 57-92%) | 19/24 (79%)<br>(95% CI: 57-92%) | >0.9 |

<sup>1</sup>n/N Non-missing (%); <sup>2</sup>Fisher's exact test; Pearson's Chi-squared test. A participant was regarded as motivated if they answered Yes with an upcoming appointment, Yes with plans to make an appointment, or if they were already regularly seeing one.

During their hospital stay, most participants reported independent toothbrushing practices (74% (118/ 158), 13.9% (22/158) required assistance and 11.4% (18/158) did not brush. Their opinions of cardiac nurses’ knowledge of oral care were mixed (Table 1). Only 21% (33/158) thought their knowledge was sufficient whilst only 34% (53/158) believed cardiac nurses could identify oral health problems. Despite this, 82.9% (131/158) of participants would consider oral health advice given by a cardiac nurse and 88.0% (139/158) would make an appointment if the nurse provided a dental referral. However, most participants 85.4% (135/158) agreed an assessment completed by a dentist during their hospital stay would be beneficial. Reflecting on their participation in CEAP, at 6 weeks, 85% (112/131) agreed that a dentist should conduct an assessment as part of their initial appointment (Figure S2 Supplementary File 5). This continued in the subset proceeding to 12-weeks.

### Study implementation

The cardiac nurses within CEAP were asked to provide anonymous feedback on the trial. All (n=7) felt it would be beneficial to patients if an oral health practitioner conducted a dental assessment during the patients’ initial CEAP appointment. Whilst one was neutral about the benefit of DOHE, all agreed that the study raised oral health awareness among both patients and staff across CEAP locations (Figure S3 Supplementary File 5).

The acceptability and perceived value of the video intervention was assessed by 83.5% (132/158) of participants. Most (≥81.9%) reported each video had educational value, were likeable and easy to understand, reinforcing the validity of their content (Figure S4 Supplementary File 5).

## Discussion

Poor oral hygiene was highly prevalent in this population with CVD attending cardiac rehabilitation and despite the prevalence of poor oral health, only 5.1% of participants reported receiving oral health education after their cardiac diagnosis.

The current study found both face-to-face and digital only oral health counselling and education significantly improved multiple markers of oral health including reduced API scores and improved oral hygiene behaviours and knowledge. Interestingly, the intervention had no significant impact on self-reported measures of participant confidence in oral self-care, participant perception of their own oral health status and participant’s motivation to visit an oral health practitioner. While face-to-face oral health education is effective, it maybe resource intensive and this study finds that the digital only intervention was also effective and may offer an initial step to a scalable solution for integrating oral health care into cardiovascular disease patient care.

The opportunity to address oral health among patients with CVD has been examined in some prior studies.^22,23^ One study of stroke in-patients involved three arms: the control arm received OHI only (similar to our study) and achieved a 40% plaque reduction, and the two intervention arms received OHI plus mouthwash and/or bi-weekly assisted brushing during their hospital stay; achieving a 68% and 74% plaque score reduction respectively.^22^ However, all study participants, including participants in the control arm, were provided with an electric toothbrush which has been shown to be more effective at plaque removal compared to a manual toothbrush.^24^ This may explain why they were able to achieve better plaque reduction compared to the participants in our study where 70% used a manual brush. Similarly in another study among stroke rehabilitation out-patients, participants receiving a 30 minute individualised oral hygiene training session and electric tooth brush achieved a greater reduction in plaque score compared to the control arm receiving the same oral hygiene training and a manual brush (43.7% vs 11.5%).^23^

Direct comparisons on the quantity of plaque reduction across studies are limited as they have varying plaque scoring methods. The two studies among participants with stroke used the Silness and Löe^25^ plaque index and the Ainamo and Bay^26^ gingival bleeding index, both of which score 6-sites around the circumference of each tooth; using either a half-mouth^22^ or full mouth protocol.^23^ The current study used the API/SBI, a modified dichotomous (yes/no) index, that records both the presence of plaque and bleeding between teeth.^27,28^ These areas are inherently more plaque retentive and prone to inflammation; commonly serving as primary sites for periodontal disease initiation.^29^ Given that patients require greater skill and attention to effectively clean in between teeth, utilising API/SBI compared to traditional indices helps to distinguish between true changes in oral hygiene behaviours and temporary improvement as a result of pre-appointment brushing^30^ and may also explain the higher reduction in plaque scores reported using traditional indices.

Despite the API reduction in the intervention groups in our study, few participants achieved optimal API (≤35%) and SBI (≤25%) scores. This may be unsurprising as oral health education likely needs to be supplemented by a professional dental clean when there is a high oral plaque burden. Compared to other effective intervention studies, similar rates of optimal API are reported. For example one comparable study, involving a population within the German Armed Forces, 9% and 19.4% of participants reaching optimal API and SBI scores respectively.^31^ However their intervention included both oral health education and a professional dental clean.^31^

Gingival bleeding is considered a marker of poor oral hygiene and inflammation as it is generally triggered by the accumulation of oral biofilm.^6^ However in the current study the mean SBI score increased over the study period, despite the improvement in oral hygiene. Similar findings were reported in patients living with stroke receiving oral health education, that is their plaque scores reduced by 40% while their gingival bleeding scores increased by 6%.^22^ It is likely that use of gingival bleeding to assess oral inflammation may be confounded in this patient population due to the high use of anticoagulant and/or anti-platelet medications;^32,33^ as well as nicotine cessation,^34^ both of which are linked to increased gingival bleeding.

The observed improvement in oral hygiene behaviours in both intervention arms (OHI+DOHE 70.5%, DOHE 55.8%) were aligned with previous studies. For example among participants attending stroke rehabilitation 23.5% adopted regular brushing (defined as ≥1 x daily) following an oral health training session.^22^ Also among cardiothoracic patients receiving individualised oral health education, their interdental cleaning habits improved in intervention and control arms by 88.6% and 45.7% respectively.^35^Importantly, their participants received weekly reinforcement of oral hygiene instructions by oral health practitioners during their stay.^35^ In contrast, our study provided a single educational intervention at baseline, which was repeated at the 6-week follow-up and was able to achieve a similar magnitude of behavioural changes. The large increase in behavioural change noted in our study may reflect the effectiveness of both face-to-face and digital education in enhancing oral hygiene behaviours in cardiac^22,23,35^ and dental^36^ settings respectively. Previous studies have not implemented digital oral health education targeting individuals living with CVD.

The baseline understanding of the impact of oral health on heart health, overall oral health knowledge, and the oral implications of heart medications was low at baseline in the current study, and is consistent with findings from an observational study conducted in a similar setting in Australia.^15^ Although over 90% of participants in the intervention groups improved in two categories: the link between oral health and CVD as well as general oral health knowledge; knowledge regarding the oral implications of heart medications remained low and is likely a result of the limited information contained within the education package. Surprisingly, those receiving DOHE alone demonstrated a better understanding of the impact of oral health on heart health, possibly due to digital media’s ability to increase health knowledge across literacy levels.^37,38^ This is the first known study to show such an improvement in oral health knowledge.

There were no differences between groups found in the current study with respect to confidence in oral self-care or self-rated oral health. However, at baseline, there was a high level of confidence and self-rated oral health (good or higher) despite the high API scores and oral disease burden. These findings were consistent with an Australian observational study that noted high confidence in self-care and oral health status in individuals living with CVD, even though nearly all respondents reported one or more oral health problem.^15^ These findings reinforce the established disconnect between individuals’ perceived oral health and objective clinical measurements.^39^ The oral disease burden reported in our study was similar to the national average for this age cohort, however the incidence of dry mouth was notably higher than the national average (48.7% vs 17.6%), likely a result of polypharmacy. This finding is particularly important as dry mouth increases the risk of oral disease.^40^ Previous studies have reported a much higher burden of oral disease with just over 90% of cardiac ward participants^35^ and 82.4% of those attending out-patient stroke rehabilitation being diagnosed with moderate-severe periodontal disease.^23^These studies used periodontal analysis^23,35^ and panoramic radiographs^35^ to confirm disease state, while our study relied solely on disease screening, potentially underestimating actual disease prevalence in this cohort.

Regular dental attendance is key to oral disease prevention and reducing inflammatory risk in this population.^3,41^ There is no universal access to dental care in Australia^42^ which explains the low number of regular dental attenders at baseline [38.6% (defined as every 3-6 months]. Despite no detectable difference between the groups, 79% of the cohort reported motivation to visit an oral health practitioner at both follow-ups; an outcome not discussed in previous oral health education studies conducted in non-dental settings.

The acceptability of the digital intervention was high, as ≥81.9% of respondents rated the videos as educational, likeable and easy to understand. As such this study has shown that oral health education is feasible in cardiac rehabilitation out-patient clinics. Specialised resourcing of calibrated oral health student practitioners was required to provide face-to-face education. The digital intervention, while less effective on its own, could be an easily implementable, cost-effective solution for delivering essential oral health messages to this patient population. This would alleviate the burden on cardiac staff, who frequently face barriers such as time constraints, lack of training, and limited resources in providing this support.^43–45^ The value of both interventions, however, suggest that the greatest therapeutic outcome would be from joint implementation especially for older adults who have a high burden of oral disease

### Strengths and limitations

The key strength of this study is that it is the first RCT to assess the effectiveness of oral health education delivered to patients with CVD in a cardiac rehabilitation setting. Secondly, the study used a combination of methods to deliver oral health education and is the first to use digital technologies. The digital oral health education package enhanced outcomes related to behaviour change and knowledge, and while the study was not powered to detect differences between DOHE alone and usual care, the findings suggest that the DOHE package could be a cost-effective, scalable solution for delivering oral health messages to this patient population.

However, the study has several limitations, firstly we required several research assistants to maintain examiner blinding between those that delivered the intervention and conducted follow-up assessments; however, their availability and study timing constraints impacted our ability to achieve our recruitment target. Next, although routine clinical measures were recorded by a single trained examiner, treatment fidelity was not assessed. Similarly, whilst a standardised protocol was used to guide the delivery of oral health education reliability and validity were not assessed. A greater proportion of males were recruited, likely a result of higher CVD prevalence in males^1^ as well as underutilisation of cardiac rehabilitation by females.^46^ Despite follow-up window extension, there was a higher-than-expected loss to in-person follow-up making the study underpowered; which may reflect de-prioritisation of oral health and/or treatment fatigue.^47^ Participants with Asian (South, East, South-East Asia) heritage were less likely to return for the primary outcome assessment; and may be reflective of cultural fatalistic views on oral health.^48,49^ Participants were also less likely to return for the primary outcome assessment if they had private health insurance that covered dental care; likely viewing oral health education in this setting as unnecessary. As the study was conducted in public hospitals in Western Sydney, primarily serving lower socio-economic populations; larger studies involving different settings may be needed to assess the broader applicability of these findings.

## Conclusion

Oral health is often overlooked in CVD management, even though the growing body of evidence underscores its significant impact on CVD outcomes. The high burden of poor oral health among patients with CVD, the relation of periodontitis and tooth loss to worse CVD outcomes, all argue for a greater focus on oral health for CVD patients.^3–5^ The progression of periodontal disease and early decay is silent.^50^ Therefore in the absence of pain many individuals may not realise they have poor oral health or that it is worsening.^6^ Integrating oral health care within cardiology settings and reinforcing the importance of regular oral health checks for patients with CVD has the potential to improve both oral and cardiovascular health outcomes. This randomised controlled trial provides evidence that digitally delivered oral health education either alone or in combination with face-to-face oral health messaging, effectively improves oral hygiene and other oral health outcomes in patients with CVD attending cardiac rehabilitation clinics.

## Data Availability

Data will be made available following assessment of a data use proposal submitted to investigators.

## Acknowledgements

We would like to thank all the cardiac nurses and research assistants for their support of the trial.

## Trial funding

CKC is supported by a NHMRC Career Development Fellowship co-sponsored by the National Heart Foundation of Australia. The post-doctoral research position (SK) and PhD scholarship (LC) were supported by donations from the Bella-Schwarz Foundation. The foundations had no role in the design of the study; in the collection, analyses, or interpretation of data; in the writing of the manuscript; or in the decision to publish the results.

## Data availability statement

Data will be made available following assessment of a data use proposal submitted to investigators.

## Author Contributions

LC, SK, AS, and CKC conceptualised the trial. LC, SK and CKC contributed equally. SM and AS contributed equally. RZ, TK and NB contributed equally.

